# Mpox Knowledge, Risk Perception, Attitudes and Willingness to Vaccinate in Colombia’s LGBTIQ+ Communities: Online Survey (CoSex)

**DOI:** 10.1101/2025.02.03.25321611

**Authors:** Cándida Diaz-Brochero, Miguel Barriga, John Fredy Ramirez, David Santiago Quevedo, Geraldine Gomez, Juliana Mateus, Natalia Nino-Machado, Juliana Cuervo-Rojas, Zulma M. Cucunubá

## Abstract

**Background:** In the 2022-23 global clade IIb mpox outbreak, Colombia was the sixth country with the most reported cases globally and the second in Latin America after Brazil. LGTBIQ+ communities, especially those with extensive sexual networks, were particularly affected.

**Objective:** We aim to characterize the knowledge, risk perceptions, attitudes, and willingness to implement preventive measures against mpox among LGBTIQ+ communities in Colombia.

**Methods:** This was an anonymized, cross-sectional, observational study based on a population-based online questionnaire using a participatory approach.

**Results:** Among 784 participants from 66 municipalities, 49.1% were aged 18–29 years, and 89.3% were male assigned at birth. A total of 73.9% identified as homosexual, while 14.9% reported never using condoms. Casual sexual partners were reported by 45.4% in the past year (median: 3; range: 1–100), and 29.3% engaged in group sex during the same period (median: 4; range: 2–100). Temporary behavioral changes to reduce mpox risk, such as limiting sexual partners and increasing condom use, were reported by 24.1%. Suspected or confirmed mpox infection was reported by 4% of participants, with 77.4% attributing potential exposure to sexual contact. Notably, 89.5% expressed willingness to receive mpox vaccination if offered.

**Conclusions:** Our findings demonstrate the importance of academic and social communities’ cross-collaboration to understand the impact of mpox infection in this population and as a basis for planning epidemic responses to future mpox outbreaks in Colombia and Latin America.

## Introduction

The 2022–2023 mpox outbreak marked an unprecedented shift in the epidemiology of the disease, with rapid global spread beyond endemic regions in Africa. Since January 1, 2022, the World Health Organization (WHO) has received reports of mpox cases from 128 Member States across all six WHO regions. By December 31, 2024, a total of 124,753 laboratory-confirmed cases, including 272 deaths, had been documented [1]. Globally, 98.7% of confirmed mpox cases during the 2022–2023 outbreak were reported among males, with the 18–44 age group being the most affected [2]. A defining feature of this outbreak was sustained person-to-person transmission, predominantly through sexual contact. In nonendemic countries, 85% of cases were linked to sexual networks involving gay, bisexual, and other men who have sex with men (MSM) [3,4].

Colombia was among the countries most affected by the global outbreak, ranking sixth overall and second in Latin America after Brazil. In a case series of 521 confirmed cases reported in Colombia between June and November 2022 [5], approximately 98% of patients identified as gay or bisexual men, 41% were living with HIV, and sexual transmission was suspected in 95% of cases. The 2015 National Demographic and Health Survey in Colombia reported that 1% of women and 1.8% of men self-identified as homosexual [6], though the actual proportion may be higher due to underreporting. Data from the 2018 National Population and Housing Census in Colombia [7] revealed that of 14,243,223 households, 48,483 (0.34%) consisted of same-sex couples (0.15% male and 0.19% female). This corresponds to a prevalence of 6.4 same-sex couple households per 1,000 Colombian couple households, one of the highest proportions in the region when compared to Argentina (3.3), Chile (2.7), Uruguay (2.3), and Brazil (1.8) [8].

To understand the dynamics of mpox transmission during this global outbreak, it is necessary to investigate the sexual behavior of persons in the age range most affected by the epidemic, especially those belonging to the LGBTIQ+ communities. On the basis of emerging scientific evidence on the sexual transmissibility of mpox [9], the WHO recommended key protective behavior to newly affected communities to reduce mpox transmission, morbidity and mortality [10]. Specific recommendations included reducing the number of sexual partners and avoiding contexts in which the risk of mpox transmission was highest [11]. Currently, the information available from Colombian and Latin American surveys on characteristics related to the sexual behavior of individuals is quite limited and outdated. Furthermore, it is likely that the data from these surveys greatly underestimate the experience of individuals at greater risk of sexually transmitted infections.

Considering the above, the objective of this study was to characterize the sexual behavior, risk perceptions, attitudes, and willingness to vaccinate against mpox among LGTBIQ+ communities in Colombia so that communication and health actions toward the control of current and future outbreaks can be tailored accordingly.

## Materials and methods

We conducted a cross-sectional online survey that was open to the entire LGBTIQ+ population (CoSex). The participants were able to complete the questionnaire from the time of its publication on May 24, 2023, to August 30, 2023. The eligibility criteria were as follows: 1) identify themselves as a person belonging to the LGBTIQ+ community; 2) were between 18 and 65 years of age; and 3) resided in Colombia at the time of application of the questionnaire. The protocol was approved by the ethics committee of the Faculty of Medicine of the Pontificia Universidad Javeriana and was carried out on 24/11/2022, act number 21/2022. We report the results following the checklist for internet E-Surveys (CHERRIES) [12]; see Supplementary file 1. Informed consent was requested from the participants to complete the questionnaire, which is available in Supplementary file 2. Participants who agreed to complete the questionnaire were provided with information on the objectives of participation, as well as the treatment of the data obtained, specifying the protection of personal data, the maintenance of confidentiality and anonymity.

The survey’s web infrastructure employs a microservice architecture, which allows for efficient management and scalability. The user interface was developed via Angular, a modern web development framework. In contrast, server-side logic, which is responsible for data processing, leverages Spring Boot for robust backend support. All the collected data were securely stored in a MySQL database [13], which was designed to protect anonymity by excluding any information that could reveal the identity of the respondents, such as personal or sensitive information. It was mandatory to answer all the questions to complete the questionnaire and save the responses. The respondents were able to review and change their answers. It was not possible to randomize the order of the questions because some of them were dependent on the answers to the previous questions, such as the number of partners according to the presence or absence of certain sexual practices. Certain questions were conditionally displayed on the basis of responses to other items to reduce the complexity of the survey. We did not collect information about IP addresses or cookies because we decided to prioritize the privacy of the participants. We did not offer any incentive to complete the survey.

The questionnaire was developed through a collaboration between the Pontificia Universidad Javeriana and Red Somos, a civil society organization that advocates for the recognition of sexual and gender diversity, in Bogota, Colombia. The questionnaire was an adaptation of the health and sexual habits survey developed by the Spanish National Institute of Statistics, which aimed to obtain relevant information to evaluate the impact of preventive policies carried out in Spain for the prevention of the spread of HIV [14]. It was divided into three sections: 1) Sociodemographic characteristics, sexual orientation, and gender identity; 2) sexual behavior, condom use, distribution of the number of regular and casual sexual partners, group and transactional sex, and history of sexually transmitted infections, among others; and 3) knowledge, risk perception and attitudes about mpox. The questionnaire and definitions used (e.g., stable partner; casual partner; transactional sex, among others) are available in Supplementary File 3.

Before the survey was launched, we conducted a pilot test with 10 people to identify potential technical flaws related to the web page, the layout of the questions or the completion of the survey and to verify that the informed consent form, the formulation of the questions, and the response options were meaningful, understandable and appropriate to the respondents and the target audience. The final self-administered questionnaire was available online, in Spanish, as a voluntary survey on a website created specifically for the project that could be accessed via computer, cell phone, or tablet. The participants were invited to participate through a multimodal strategy that included the following: 1) social networks (Facebook, Instagram, and Twitter); 2) a contact list of LGBTIQ+ community-based organizations or organizations working in the prevention and/or care of HIV/AIDS; and 3) promotional ads on contact platforms and meetings of persons from LGBTIQ+ sectors, such as Grindr; and 4) availability of the questionnaire in places of homosocialization, such as dark rooms and saunas.

For the sample size calculation, an exploratory analysis was carried out to identify clusters of sexual behavior for which there were no previous estimates. According to Floyd & Willman, for this type of exploratory analysis, having ten individuals for each variable examined is suggested [15]. Considering a total of 73 variables, we estimated a minimum sample size of 730 participants. Nonprobabilistic snowball sampling was carried out. This technique allowed the sample size to increase as the individuals who filled out the questionnaire invited their acquaintances who met the eligibility criteria of the study to participate.

Descriptive statistical methods were used to present the results. Absolute and relative frequencies were used to describe qualitative variables. For quantitative variables, measures of central tendency and variability were calculated. The means and standard deviations are presented for variables with a normal distribution, and the medians, ranges and 25th to 75th percentiles are presented for variables with a nonnormal distribution. Normal distribution was assessed via the KolmogorovlJSmirnov test at the 5% significance level (p < 0.05). For the analysis of the reasons for not receiving mpox vaccination, we categorized and codified the information and represented it in a conceptual scheme. All analyses were performed via R software (version 4.4.1; R Foundation for Statistical Computing) [16].

## Results

There were 6494 entries into the survey, 968 of which were completed. Of these, 184 (19%) did not meet the inclusion criteria, leaving a total of 784 participants for analysis. Because we chose to analyze only information from participants who completed the entire survey, there were no missing data (Figure 1). The median age of the respondents was 30.2 years (IQR 24.8--36.6); 89.3% (n=700) reported that their sex assigned at birth was male, 83.5% (n=654) identified as male, and 73.9% (n=579) were homosexual (Table 1).

**Figure 1:**
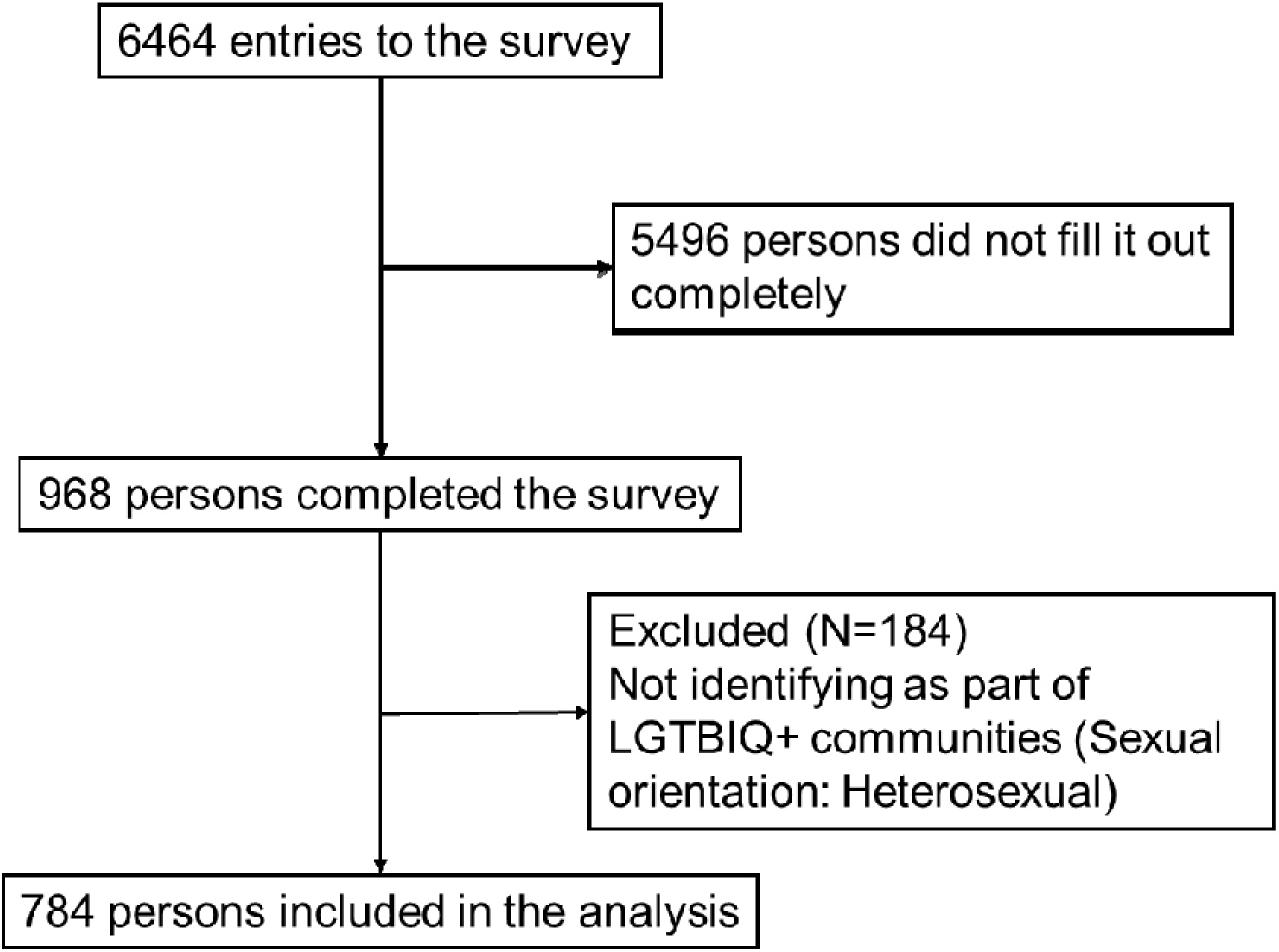
Study flowchart.

**Table 1:**
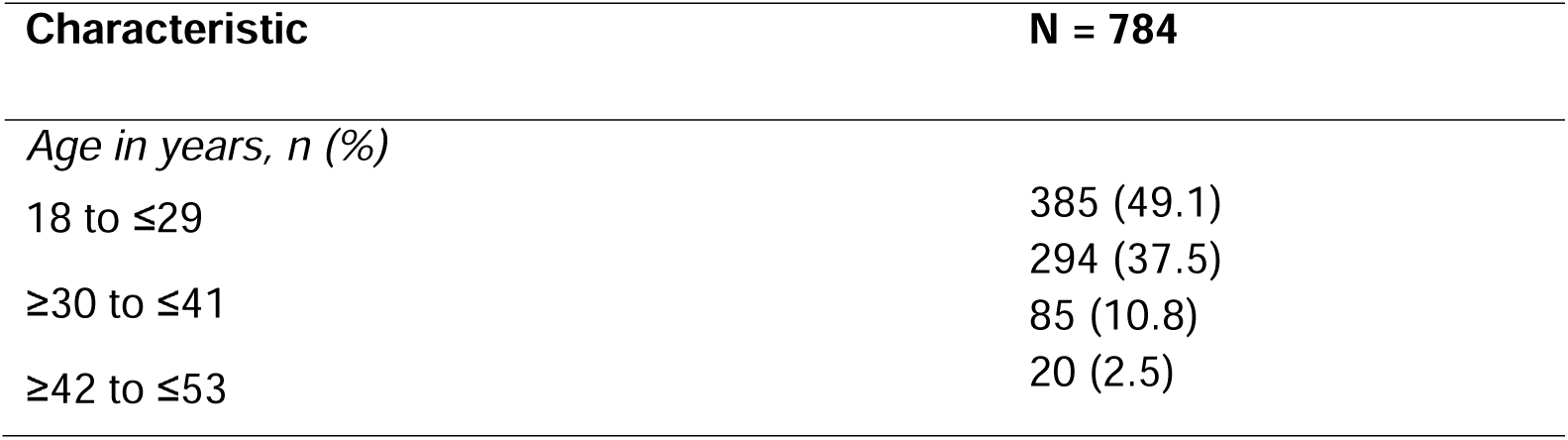
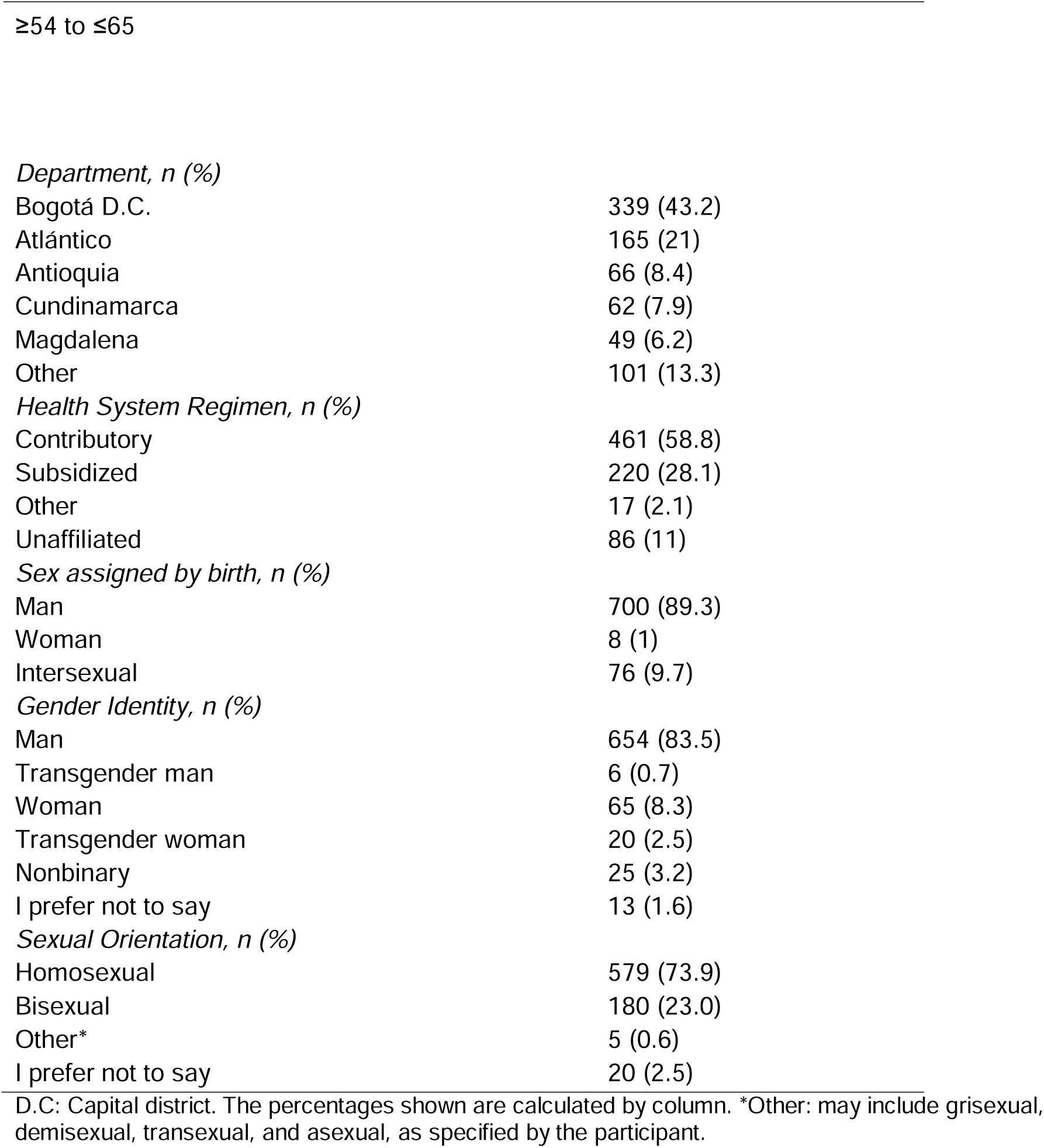
Sociodemographic characteristics, sexual orientation, and gender identity (N = 784). CoSex survey. May 24 to August 30, 2023.

In terms of respondents’ sexual behavior, the median number of sexual partners in the past year was 3 (range 0--200), 33.5% reported having one or more stable sexual partners in the past year, with a median of 1 (range 1--5), and 45.4% reported having casual sexual partners in the past year, with a median of 3 (range 1--100). A total of 14.9% reported never using condoms, and 41.9% reported sometimes using condoms. A total of 29.3% had engaged in group sex in the past year, with a median number of partners of 4 (range 2--100). A total of 25.4% reported either giving or receiving money or gifts from someone in exchange for sex. A history of diagnosed STIs was not uncommon, and the most frequently reported were syphilis, HIV, and gonorrhea infection. A total of 12.9% had received preexposure prophylaxis (PrEP) in the last month (Table 3). Four percent of the respondents reported suspected or confirmed exposure to mpox, 77.4% of whom believed that the source of infection could have been sexual contact (Table 5).

Knowledge of mpox was reported by 64% of the respondents. Knowledge about mpox-related symptoms and sources of transmission of mpox among persons who reported knowing about the disease is presented in Figure 2. Knowledge was greater among those between 42 and 53 years of age (84.7%), females assigned by birth (87.5%), those identified as transgender (83.0%), those identified as homosexual (66.3%), those residing in Antioquia (72.7%), and those belonging to the contributory health system (71.4%) (Table 2). The main sources used by the respondents to learn about mpox were social media (31.1%) and websites of community-based organizations (30.9%) (Table 4).

**Figure 2:**
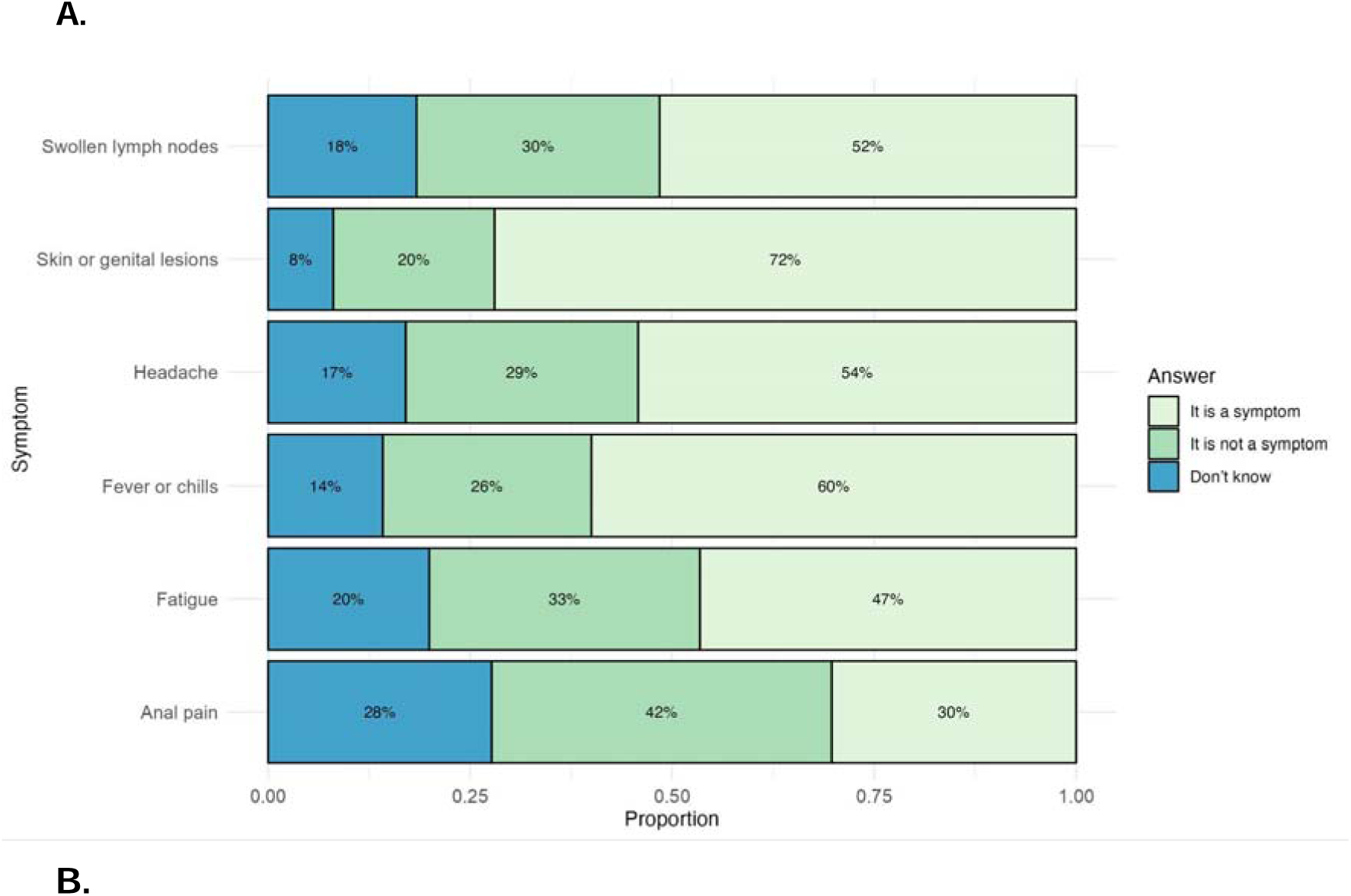
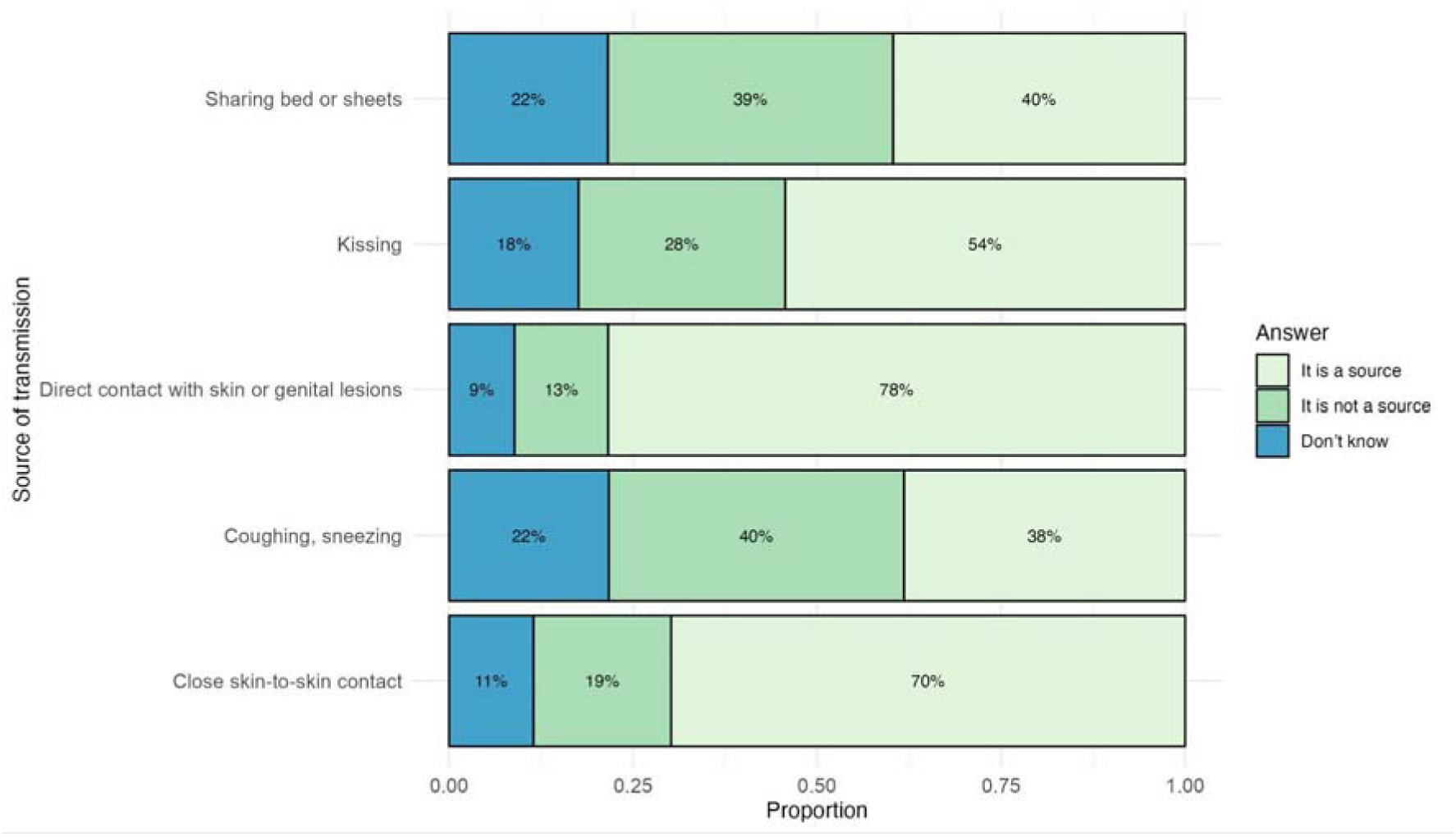
Knowledge about mpox-related symptoms **(Panel A)** and sources of transmission of mpox **(Panel B)** among persons who reported knowing about the disease (N = 502)

**Table 2:**
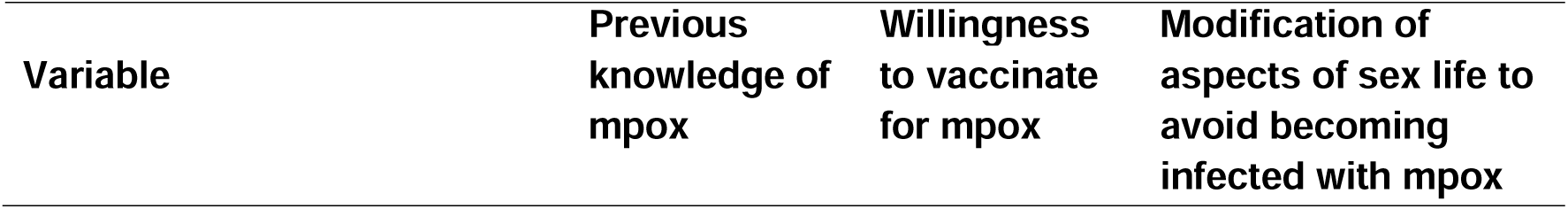
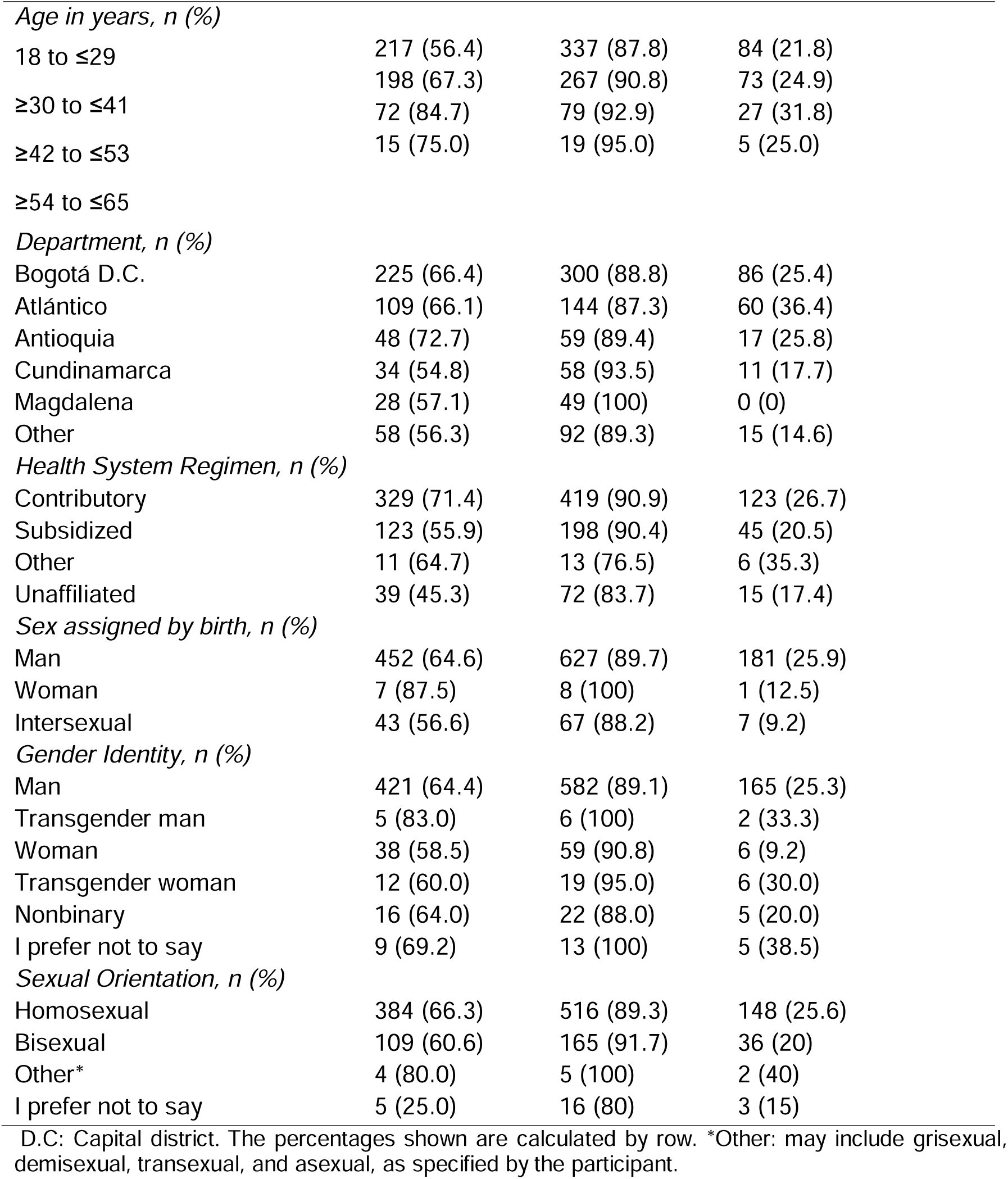
Knowledge of mpox, willingness to vaccinate and modification of aspects of sexual life to avoid becoming infected with mpox, according to sociodemographic characteristics, sexual orientation, and gender identity (N = 784). CoSex survey. May 24 to August 30, 2023.

**Table 3:** Sexual behavior and practices of the respondents (N = 784). CoSex survey. May 24 to August 30, 2023.

| **Behavior and practices** | **N=784** |
| --- | --- |
| Number of sexual partners in the last year, median (Range; 25th-75th percentile) | 3 (0-200; 1.5-10) |
| Number of sexual partners in the last month, median (Range; 25th-75th percentile) | 1 (0-45; 1-3) |
| *Means to meet new sexual partners\*, n (%)* |  |
| Dating applications (Grinder, Scruff, others). | 467 (59.6) |
| Social networks (Facebook, Twitter, Instagram or TikTok). | 407 (51.9) |
| Through friends or acquaintances. | 315 (40.2) |
| Homosocialization or flirting sites (dark rooms, saunas, bars. | 227 (28.9) |
| Cruising sites (public restrooms, parks) | 155 (19.7) |
| Mass gatherings (parties, orgies, others) | 131 (16.7) |
| Have stable sexual partner(s), n (%) | 263 (33.5) |
| Number of stable sexual partners, median (Range; 25th-75th percentile) | 1 (1-5; 1-1) |
| *Condom use in the last year, n (%)* |  |
| Always | 344 (43.9) |
| Sometimes | 323 (41.2) |
| Never | 117 (14.9) |
| Condom use at last intercourse, n (%) | 456 (58.2) |
| Participation in group sex in the past year, n (%) | 230 (29.3) |
| Number of sexual partners in group sex, median (Range; 25th-75th percentile) | 4 (2-100; 3-6) |
| Having casual sexual partner(s) in the past year, n (%) | 356 (45.4) |
| Number of casual sexual partners in the past year, median (Range; 25th-75th percentile) | 3 (1-100; 2-9) |
| Giving money or gifts to someone for having sex in the past year, n (%) | 87 (11.1) |
| Number of partners to whom you have given money or gifts for having sex in the past year, median (Range; 25th-75th percentile) | 2 (1-30; 1-3) |
| Receiving money or gifts for having sex in the past year, n (%) | 112 (14.3) |
| Number of partners from whom you have received money or gifts for having sex in the past year, median (Range; 25th-75th percentile) | 3 (1-102; 1-5) |
| *Diagnosis of one or more of the following sexually transmitted infections in the past\*, n (%)* |  |
| Syphilis | 209 (26.6) |
| HIV | 143 (18.8) |
| *Neisseria gonorrhea* | 69 (8.8) |
| Genital herpes | 67 (8.5) |
| Condylomas or genital warts | 62 (7.9) |
| Sexually transmitted infection (but not told or do not remember which) | 37 (4.7) |
| Hepatitis B | 28 (3.6) |
| *Chlamydia trachomatis* | 25 (3.2) |
| Hepatitis C | 12 (1.5) |
| Receiving PrEP for HIV prevention in the last month, n (%) | 101 (12.9) |
| *Means of receiving PrEP<sup>a</sup>, n (%)* |  |
| Health insurance company | 79 (78.2) |
| Nongovernmental organizations | 12 (11.8) |
| Friends or acquaintances | 5 (4.9) |
| Homosocialization or flirting sites (orgies or chem sex encounters) | 5 (4.9) |
| Have received PEP for HIV prevention in the past year, n (%) | 82 (10.5) |
| *Means of receiving PEP for HIV<sup>b</sup>, n (%)* |  |
| Health insurance company | 62 (76.5) |
| Nongovernmental organizations | 14 (17.3) |
| Friends or acquaintances | 5 (6.2) |
| *Nonmutually exclusive categories. <sup>a</sup> Calculated over 101 participants who received PrEP. <sup>b</sup> Calculated over 82 participants who received PEP. HIV: Human immunodeficiency virus; PrEP: preexposure prophylaxis for HIV; PEP: postexposure prophylaxis for HIV. |  |

**Table 4:** Knowledge of mpox and sources used by respondents to learn about mpox (N = 784). CoSex survey. May 24 to August 30, 2023.

| Characteristic | N = 784 |
| --- | --- |
| Know or have heard about mpox, n (%) | 502 (64) |
| *Mpox information resources\*, n (%)* |  |
| Social media | 244 (31.1) |
| Websites of community-based organizations | 242 (30.9) |
| Newscasts/Television programs/Newspaper/Radio | 201 (25.6) |
| Official media of WHO or PAHO. | 158 (20.1) |
| Colombian National Institute of Health, Ministry of Health of Colombia | 139 (17.7) |
| Family or friends | 80 (10.2) |
| *Nonmutually exclusive categories. WHO: World Health Organization; PAHO: Pan American Health Organization |  |

**Table 5:**
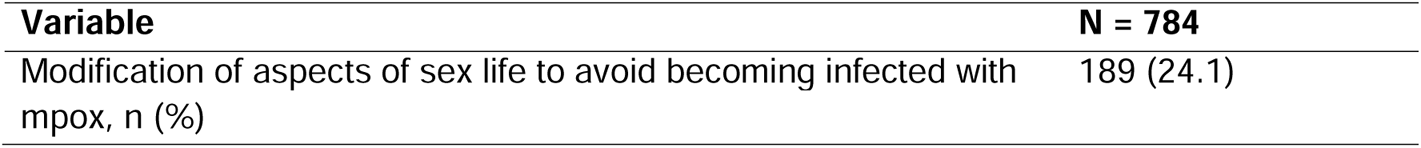
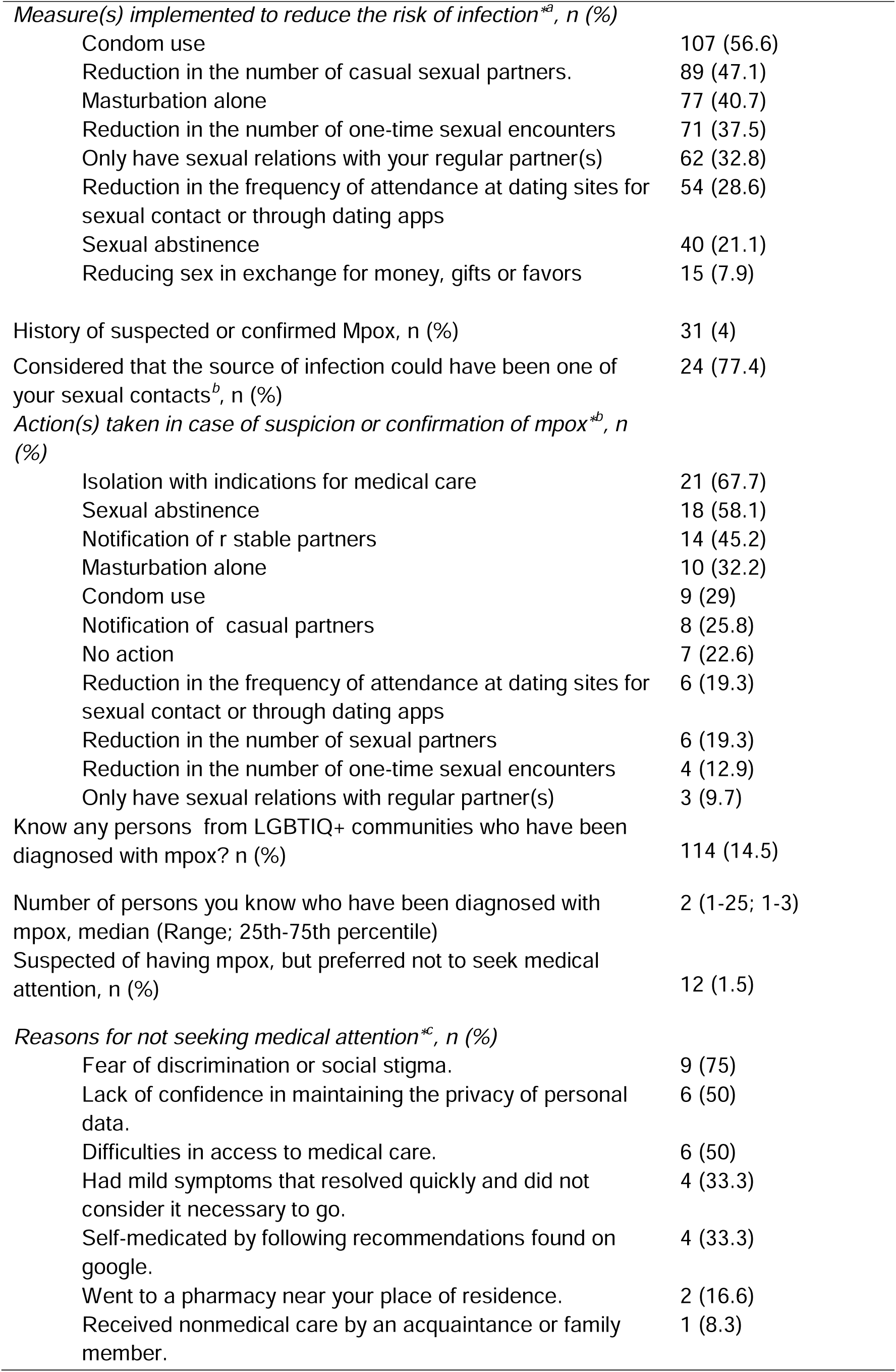
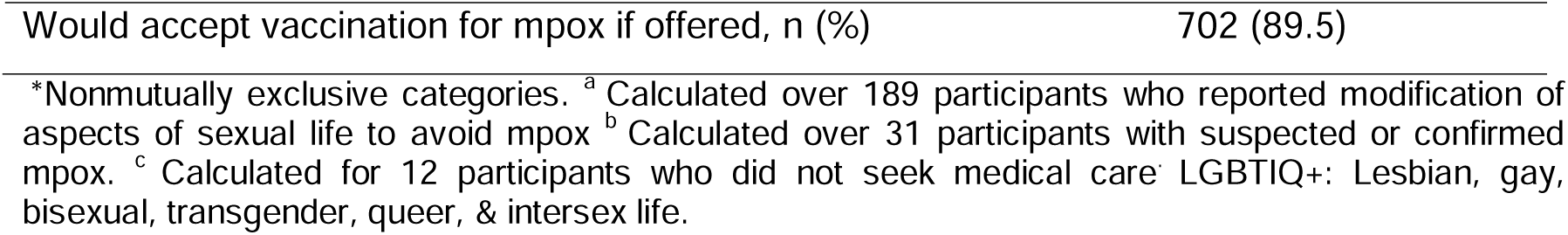
Risk perception and attitudes toward mpox (N = 784). CoSex survey. May 24 to August 30, 2023.

Modifications of aspects of sex life to avoid becoming infected with mpox were reported in 24% of the respondents. The persons who most frequently reported having modified their sex lifestyles to avoid contracting the virus were those between the ages of 42 and 53 years (31.8%), sex assigned at birth (25.9%), homosexual (25.6%), transgender (33.3%), and residing in Atlántico (36.4%) (Table 2). The main measures implemented to reduce the risk of mpox infection were condom use (56.6%) and reducing the number of sexual partners (47.1%) (Figure 3). A total of 1.5% suspected that they had mpox but chose not to seek medical attention, with 75% citing fear of discrimination or social stigma as the main reason (Table 5 and Figure 4).

**Figure 3:**
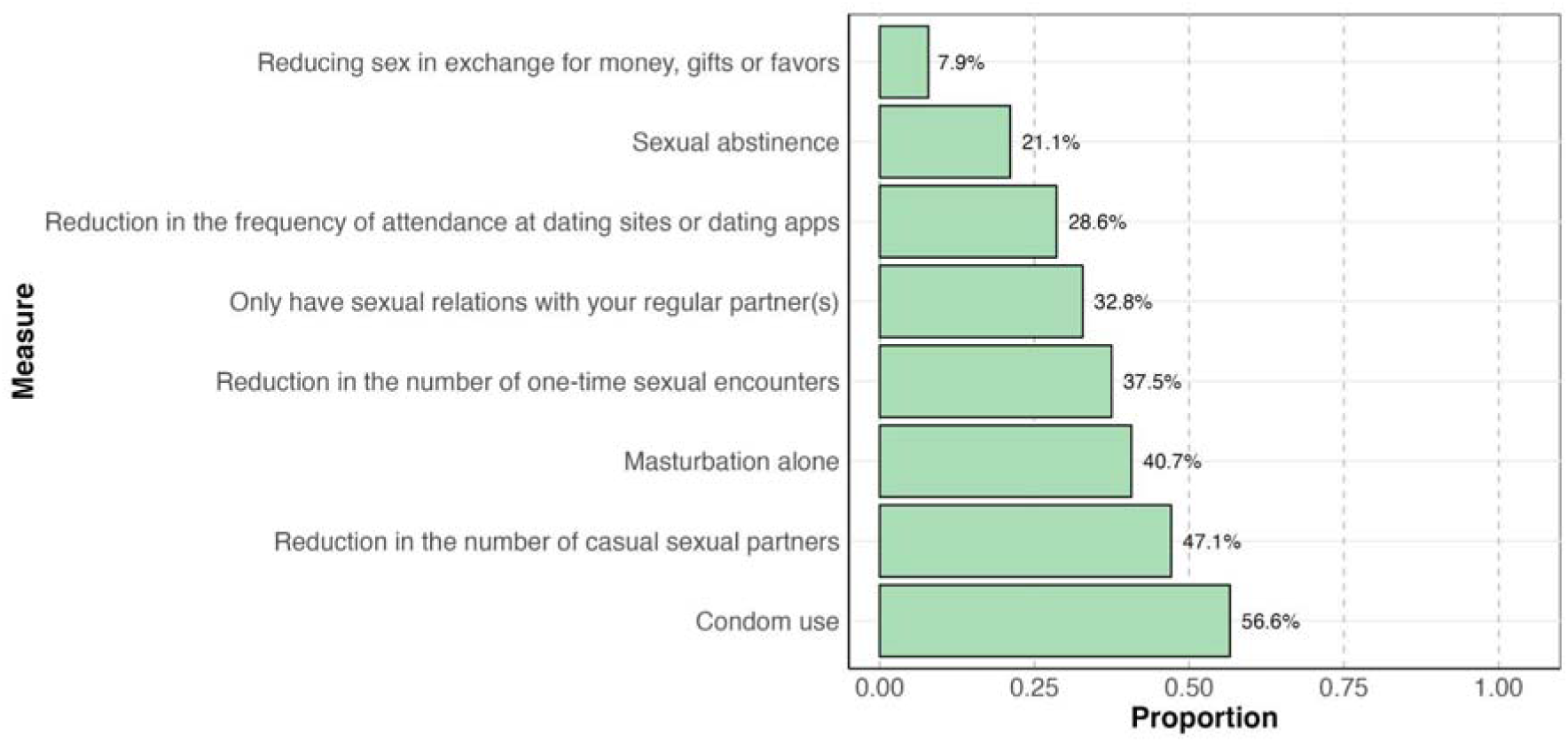
Measure(s) implemented to reduce the risk of mpox infection (N=784)

**Figure 4:**
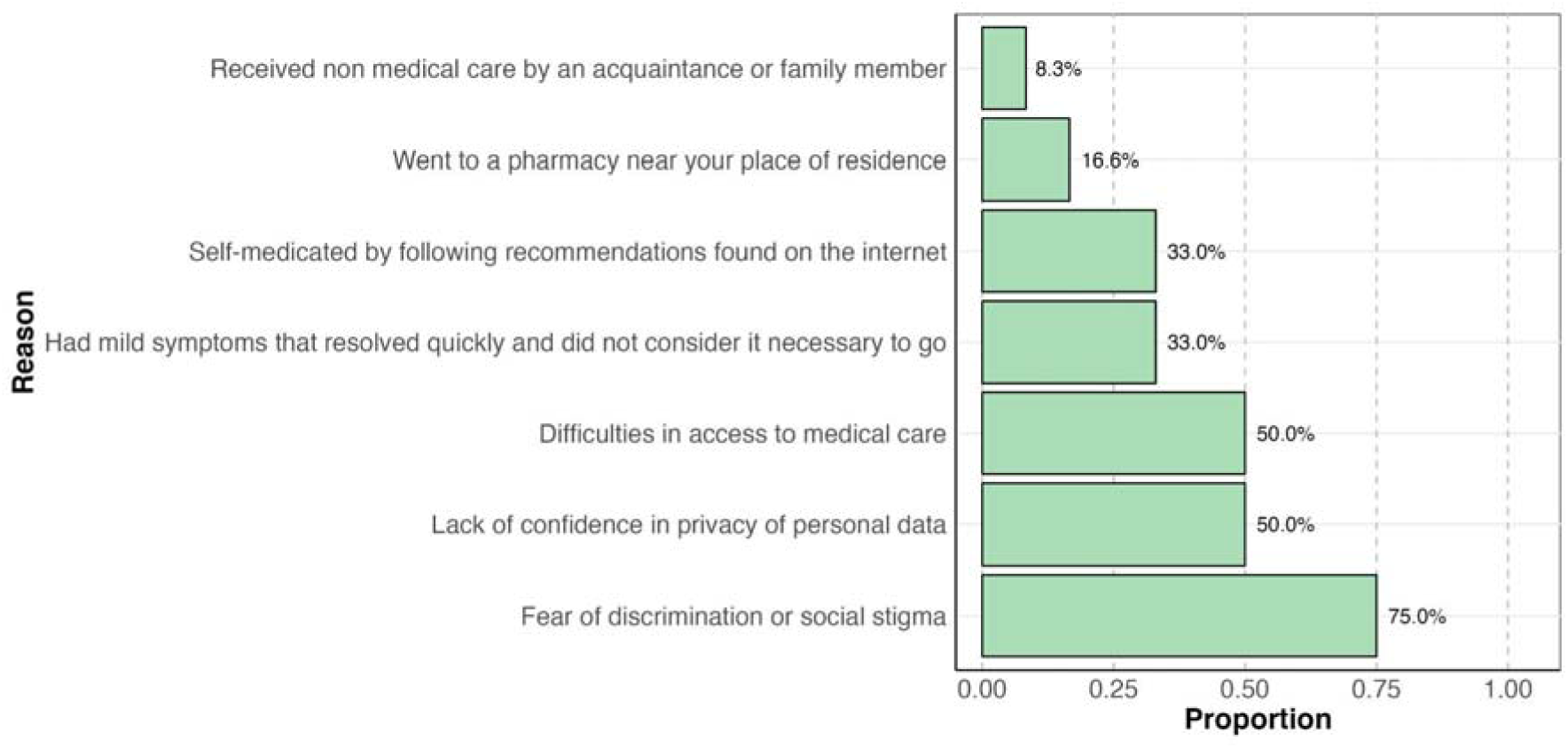
Reasons for not seeking medical attention in cases of confirmed or suspected mpox disease (N=12)

Willingness to be vaccinated was reported by 89.5% of the respondents. It was higher in those aged 54--65 years (95%), those with sex assigned at birth (87.5%), those identified as transgender men (100%), those identified as bisexual (91.7%), those residing in Magdalena (100%), and those belonging to the contributory health system (90.9%) (Table 2). The reasons for not receiving the vaccine are described in Figure 5.

**Figure 5:**
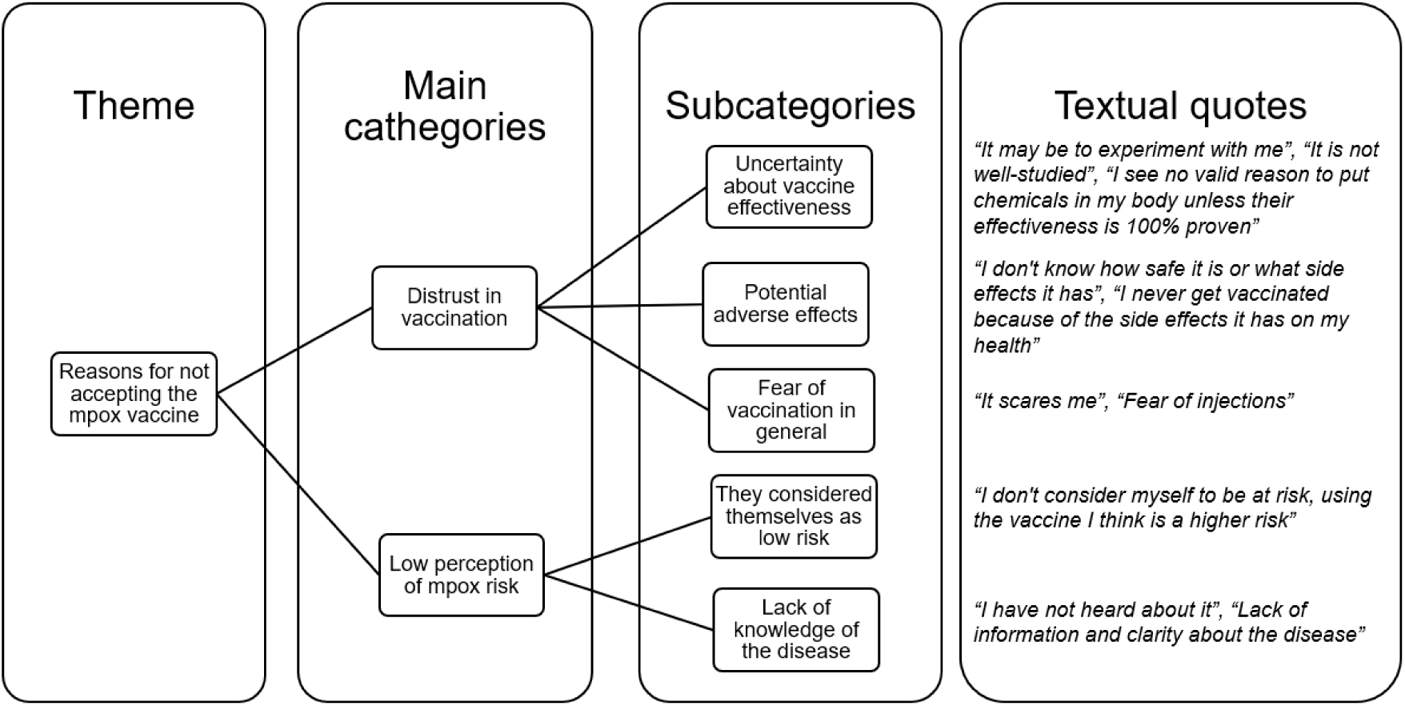
Categorization of the reasons for not accepting the mpox vaccine if it was offered to them (N=82)

## Discussion

### Principal findings

We present a descriptive analysis of the results of an online-based survey in Colombia among LGTBIQ+ communities conducted in 2023 during the mpox outbreak acceleration phase of the epidemic in the country. The respondents were predominantly from metropolitan areas; 89.3% were male, 86.6% were between 18 and 41 years of age, and 73.9% were homosexual. A history of suspected or confirmed mpox in the survey was reported by 4%, which is significantly higher than the country’s surveillance data at the time [17], suggesting that we may have reached a population at higher risk of infection. We found generally poor to moderate knowledge of the symptoms and sources of transmission of mpox. Only 24% of the respondents said that they had changed aspects of their sex life to avoid infection. Close to 90% would be willing to be vaccinated if a vaccine is offered, with fear and lack of knowledge about the vaccine being the main reasons for nonacceptance.

### Knowledge of the symptoms and sources of transmission of mpox

The survey was conducted at a time when information about the risk of contracting mpox had been disseminated in the mass media and some public health campaigns aimed primarily at the MSM population. Despite this, regarding prior knowledge of mpox, we found that only 64% reported having previously heard of the disease. Among those, 38% to 78% correctly identified various possible transmission sources, and 30% to 72% correctly identified various symptoms associated with mpox, demonstrating moderate knowledge of the disease. These findings are similar to those reported in a qualitative study in the U.S. [18], where only 35% of respondents were classified as having good knowledge of the disease, defined as answering at least 60% of the questions correctly. In addition, our results showed that the main source used by respondents to learn about mpox was social media, which is a concern given the high rate of false, misleading and harmful information that has been shown to circulate on these platforms [19]. This also highlights the importance of early and clear information from official institutions, such as the WHO, to the most affected communities, with a nonstigmatizing message.

### Sexual behavior and practices among the respondents

In the setting of epidemics with a potential sexual mode of transmission, such as mpox, the degree of sexual activity of susceptible individuals exerts a preponderant influence on the transmission of the infection, such that individuals with the greatest number of partners bear the greatest burden of disease [20]. Our results revealed that only 33% reported having stable partners, 56.1% reported either never using or sometimes using condoms during sex, 29% participated in group sex, 14% received money or gifts for having sex, 45% had casual or one-time partners in the previous year, 26.6% reported having been diagnosed with syphilis in the past, and 18.8% reported living with HIV. These variables are related to a greater number of sexual contacts that may have an impact on the likelihood that an infection introduced into the sexual network will lead to an outbreak, as well as on the size and duration of the outbreak [21].

### Risk perception and attitudes toward mpox

With respect to risk perceptions and attitudes toward mpox, we found a surprisingly low frequency of modification of sexual behavior to prevent mpox (24%) compared with data from a survey in the U.S., where 83.5% of participants had adopted at least one sexual risk-reduction behavior to avoid mpox infection [22], and compared with 50.9% in another large survey in 23 countries in Europe and the Americas [23], reflecting a relatively low-risk perception among respondents. This could be partly explained by the timing of the survey (after the initial peak of the epidemic), when the number of cases in the country and around the world was decreasing but also by a lack of knowledge and awareness of the disease and possibly recall bias. The main measures taken to prevent mpox were condom use, a reduction in the number of casual sexual partners and masturbation alone, which is consistent with other similar surveys in Brazil [24], Australia [25] and the United States [26]. According to the results of a regression model by Prochazka et al. [23], individuals who reported concern about mpox were more likely to adjust their sexual behavior, whereas participants who reported vaccination or had a history of mpox were less likely to continue adjusting.

Our results show that only 1.5% of the respondents did not seek medical care upon suspecting that they had been afraid. This number is very low, compared with the 96% reported in a similar study in Brazil [24]. The three main reasons for not seeking medical attention were fear of discrimination or social stigma, lack of confidence in maintaining the privacy of personal data and difficulties in accessing medical care.

This highlights the importance of optimizing communication strategies among both health workers and communities and generating more expeditious medical care routes in the context of an epidemic.

### Willingness to accept vaccination and other preventive measures

Our results revealed a high willingness to vaccinate against mpox in Colombia (89.5%). Recently, a systematic review reported an acceptance rate of mpox vaccination of 73.6% among the LGBTIQ+ communities globally and 60.9% in the Region of the Americas [27]. Among the reasons for nonacceptance in our study, we found a lack of confidence in the efficacy of the mpox vaccines and vaccines in general, fears and doubts about experimental studies, concerns about vaccine safety and potential adverse effects, and a low perception of the risk of infection. Recent studies have attempted to predict factors associated with the mpox vaccine among MSM and sexual minorities. Individuals with higher levels of fear of social rejection due to mpox acquisition [28], fear of mpox [29], being nonheterosexual [29], having socioeconomic instability [28], living in rural areas [30], not knowing mpox vaccinated persons [30] and hesitant about the vaccine [28] are more likely to reject the vaccine or be unvaccinated.

Importantly, although Colombia was the six country most affected by the mpox outbreak in the world and the second most affected country in Latin America after Brazil [17], vaccination in the target high-risk population only started in January 2024 in the context of a clinical trial to evaluate the live attenuated vaccine LC16m8 [31]. Other Latin American countries, including Chile, Brazil and Peru, had previously initiated vaccination plans as part of their national strategic immunization programs, which targeted priority groups with the live, nonreplicating JYNNEOS® vaccine, which was approved by the FDA for mpox prevention [32].

### Strengths and limitations

A major strength of this study is that it was able to collect important information on knowledge, sexual behavior, risk perceptions and attitudes toward mpox in LBTGQ+ communities, analyzing one of the largest samples recruited to date in Latin America. Another important strength is that the questionnaire design and recruitment were carried out in collaboration with one of the largest community-based MSM organizations in Colombia, using a methodology aimed at avoiding forms of discrimination or stigmatization in LGTBIQ+ communities.

However, it is important to acknowledge several limitations: 1) The data came from a subset of participants, mainly from metropolitan areas, which may present different risks. The representativeness of all the LGTBIQ+ persons in Colombia is not known; thus, the results cannot be generalized to all the LGTBIQ+ communities in the country. 2) All of the data were self-reported and are potentially subject to social desirability bias, although this bias is likely to have been minimized because of the self-administered anonymous nature of the survey. 3) To protect the privacy and confidentiality of the information, we did not collect information about IP addresses or cookies to identify unique visitors; thus, we were not able to calculate the view or participation rate. Nevertheless, we assumed that the completed attempts to answer the survey corresponded to individual subjects, since the response patterns are different, and it is unlikely that an individual would answer the survey more than once.

## Conclusions

This study provides valuable insights into sexual behaviors, risk perceptions, and attitudes toward mpox among LGBTIQ+ communities in Colombia during the mpox outbreak. While knowledge of mpox symptoms and transmission was moderate, many respondents engaged in high-risk sexual behaviors, including having multiple casual partners, participating in group sex, and inconsistent condom use, among others, which could facilitate mpox transmission. Although most participants were willing to be vaccinated, challenges such as fear of stigma, doubts about vaccine efficacy, and low perceptions of personal risk need to be considered. Our findings demonstrate the importance of academic and social communities’ cross-collaboration to understand the impact of mpox infection in this population and as a basis for planning current and future epidemic responses. Additionally, our results highlight the need for targeted public health interventions, including tailored education campaigns and accessible vaccination strategies, to address the specific needs and concerns of these communities.

## Data Availability

All data produced in the present study are available upon reasonable request to the authors

## Abbreviations

WHO: World Health Organization
HIV: Human immunodeficiency virus
LGBTIQ+: Lesbian, gay, bisexual, transgender, queer, intersex, plus
MSM: men who have sex with men
PrEP: preexposure prophylaxis
PEP: postexposure prophylaxis
STI: sexually transmitted infection

## Declarations

No conflicts of interest reported

## Availability of data and materials

The data and R code used for the statistical analysis are publicly available at https://github.com/TRACE-LAC/CoSex.

## Author contributions

CDB, ZMC, DSQ, MB, and JFR conceived this study. CDB and DSQ drafted the questionnaire. ZMC, MG, JFR, JCR, GG, JM, and NN reviewed the questionnaire. MB and JFR conducted the field work. CDB, DSQ, JCR, and NN conceived and supervised the current analysis. GG supervised data acquisition online. CDB and DSQ performed the statistical analysis. CDB and ZMC drafted the manuscript. CDB, ZMC, DSQ, MB, JFR, JCR, and NN revised the manuscript for important intellectual content. All the authors read and approved the final manuscript.

## Ethical Approval Statement

The protocol was approved by the ethics committee of the Faculty of Medicine of the Pontificia Universidad Javeriana, carried out on 24/11/2022, act number 21/2022.

